# Summative evaluation of the Rural Surgical Obstetrical Networks Initiative: Findings from a five year retrospective qualitative study

**DOI:** 10.1101/2025.09.29.25336918

**Authors:** Jude Kornelsen, Audrey Cameron, Tom Skinner

## Abstract

**Background:** The Rural Surgical and Obstetrical Network (RSON) initiative was originally funded for five years to support, enhance, and sustain rural surgical and obstetrical services in British Columbia (Canada). Interviews and focus groups with healthcare providers and administrators from 10 rural communities were conducted annually, to document the implementation of RSON, understand challenges and gains, and to inform network improvements. To supplement this information and enable participants to reflect on the project’s full duration, we conducted a summative evaluation of RSON to understand the extent to which the program achieved its objectives.

**Methods:** Data were collected in two phases: annual interviews and focus groups with rural healthcare providers and administrators in the final project year and through a real-time interactive, anonymous platform, ThoughtExchange, in the context of a day-long, annual virtual meeting. Data from both sources were analysed using a process of thematic analysis.

**Findings:** Across both data sources, common themes highlighted the benefits of RSON, including the stabilization of local procedural care through increased local scope and volume, support for clinical coaching and other ongoing educational initiatives, quality improvement efforts, team-oriented care, increased administrative support, and appropriate staffing levels. Themes unique to the summative interviews included those addressing the limitations of RSON funding, particularly constraints on funding and contextual factors limiting participation.

**Conclusion:** Findings underscore the importance of a wrap-around approach to health service interventions to reflect the complexity of healthcare delivery and the importance of coordinated, context sensitive efforts that work in synergy with other initiatives. The RSON initiative precipitated a cultural change in key determinants of stability at individual site levels, a necessary precursor to long-lasting change. The combination of this retrospective assessment of RSON and the program evaluation findings suggest the need for long-term funding to support rural surgical and obstetrical services.

## Introduction

Clinical networks of care have gained in popularity as a solution to healthcare organization across interprofessional domains and expansive geography, the latter particularly relevant to rural and remote health services. They can include a broad range of healthcare providers, social service organizations, and patients, with the specific configurations responding to the objectives of the network (1). Benefits to network models include improved access to care due to established pathways to specialist care for those from rural sites and enhanced coordination of care between providers (2). Efficiencies gained through enhanced coordination may yield spinoff effects, including reduced health system costs due the elimination of duplicated services and a more effective use of resources. This may lead to better recruitment and retention of healthcare providers due to the increased local healthcare stability (3).

The Rural Surgical and Obstetrical Networks (RSON) initiative in British Columbia was funded by the Joint Standing Committee on Rural Issues to stabilize and grow low-volume surgical and obstetrical services that were at risk of closure. The approach was driven by the need to establish supportive structures (“pillars”) that improve service sustainability, including 1) funding for resources to increased scope and volume at local sites; 2) support for wrap-around continuous quality improvement, 3) initiation of clinical coaching programs for care providers, 4) use of remote presence technology to connect care providers with regional specialists, and 5), comprehensive, ongoing evaluation. As a part of the evaluation, both baseline data and annual data were gathered to assess the efficacy of these pillars and the network structure. The lead author contributed to the conceptual development of the networked surgical services model and created the strategic approach to a comprehensive, mixed-methods evaluation, informed by co-development of evaluation methods with key partners. Findings from other sub-studies within the RSON evaluation demonstrated positive effects on process at local levels through improved team function (4), improved regional team function (5), patient satisfaction with services (6), positive effects of continuous quality improvement (7), and increased understanding of site capacity by regional specialists (8). Clinical findings for low acuity populations report health outcomes comparable to those at higher volume, specialist-led sites for obstetrical care (9), and comparable outcomes based on provider type (specialist or generalist) (10).

In this paper, we summatively identify the overarching “wins” of the initiative from the perspective of care providers and administrators. We anticipate our findings will be useful for future planning of rural clinical networks, both within local jurisdictions and across regions.

## Background

The importance of evaluating any health system intervention is well established (11,12). At the highest level, evaluation enables an assessment of the *value* of public investment against the *costs* of such investment. Evaluating effectiveness allows for robust policy and decision-making regarding continued investment, implementation in other jurisdictions, and the identification of program weaknesses and corrective measures. Ultimately, a robust, comprehensive evaluation holds the implementers accountable to the use of allocated funds, ensuring effective, efficient, and ethical use.

The RSON initiative recognized the importance of such an evaluation and embedded it in the structure of the network rollout. This was achieved primarily through a mixed-methods, ongoing program evaluation, including annual site visits to participating communities, targeted quantitative surveys (for example, to outreach specialists and patients who engaged in virtual pre-and post-procedural care), and analysis of administrative data to understand health outcomes. A Delphi process was used to ensure the evaluation measures reflected participant priorities (13). These initiatives allowed for the collection of multiple datasets to support program evaluation (14). The evaluation was informed by existing frameworks (15–17), and key partner engagement (18), in addition to reviewing existing literature. Further, we aimed to focus outputs on care providers and policy and decision-makers to ensure health system relevance (19).

Network evaluations are notoriously difficult, due, in part, to their amorphous, organically evolving nature (20,21). Given this, most network evaluations use a mixed-methods approach to combine the richness of qualitative insight with more quantitative measures gained through surveys, interviews, and document reviews (22). Such findings can serve as useful indicators for scale and spread to other jurisdictions.

Many network evaluations focus specifically on the internal structure of the network itself. For example, Cunningham et al. (2019) focused their evaluation at the community, network, and organizational level (23). Others focus on the relationship between network architecture and outcomes, and consider evaluation frameworks from the perspective of “pillars”, including “network connectivity” (membership and organization/structure of network), “network health” (resources and infrastructure for sustainability, and the systems that the network is built on), “network results” (interim outcomes as the project works towards an ultimate goal), and endpoint impact (24).

Others focus on various network attributes such as governance and leadership (23,25,26), network objectives and outcomes (25,26) (e.g. has it improved care coordination (26), program sustainability, potential for further implementation (26,27), and establishing recommendations and evidence that can facilitate continued support and funding (15,24,28).

There is limited literature on summative evaluations of rural surgical and obstetric service networks in high income countries with rural regions. An evaluation of an initiative similar to BC’s RSON program in the United States, “Rural Maternity and Obstetrics Management Strategies (RMOMS)” assessed the networks’ impact on maternal health outcomes to determine the efficacy of (and thus continued investment in) this model of care (29).

Other studies have documented evaluations of cancer care networks (30), pain management (31), telehealth (27), cardiac services (32), dental care (33) and primary care (26). Like our study, many used a qualitative approach (26,27,34) and collected data through interviews and questionnaires (26,27,31). A few studies collected quantitative data (30,32,33), one through surveys (33), the other through electronic health records (30).

In the context of rural communities, this evaluation sought to make explicit what has often been assumed: that effective rural healthcare is contingent on the strengths and functionality of frequently informal networks, operating across vast geography and often isolated communities. While networks have long been present in rural healthcare, there has been little conscious attention to evaluating how they function, or the specific enablers and antecedents that support successful network function. With this evaluation, we strove to explicate mechanisms of relationship-building and infrastructural support across network levels, between providers, administration, and regional partners, without simply assuming their presence. Before we can understand high-level outcomes such as patient outcomes, we must first examine the relational structure that underpin rural healthcare function.

To accomplish this, an iterative program evaluation was threaded throughout the course of implementation, findings from which are reported elsewhere (4–6,35–38). Adjunctively, we undertook a summative evaluation to capture “wins” and the most crucial legacy aspects of the model. This paper focuses on the summative evaluation.

## Methods and approach

These data arise from two collection phases: the first phase was through individual and focus group interviews with rural healthcare providers and administrators in the final year of data collection (January-May 2022), conducted in accordance with the University of British Columbia’s Behavioural Research Ethics board, through which approval was granted (ID: H18-01940). The second phase included the collection of participant feedback through a real-time interactive platform, *ThoughtExchange*, in the context of a day-long, annual virtual meeting (November 17, 2022).

### Interview and focus group methods

Participants were recruited during the last year of the project through third party recruitment by the Local Community Coordinator (LCC) at each site, a position supported through RSON funding. Inclusion criteria included all healthcare providers, extended team members and administrators (Family Physicians with Enhanced or Obstetrical Surgical Skills [FP-ESS/OSS] and Family Practice Anesthetists, nurses, midwives, health services administrators, local care coordinators, operating room managers and booking clerks). Participation was entirely voluntary, and no participants requested to drop out over the course of the study.

Individual and focus group interviews were conducted either in-person in hospital board rooms or virtually, using the Zoom videoconferencing platform. All were led by the principal investigator (JK), who is skilled in qualitative interviewing and has over two decades worth of experience in rural health research and evaluation. Interviews were also attended by a research assistant for fieldnote taking. The research team had a longitudinal relationship with interview participants, and participants were introduced to the overall scope of the study before beginning their interviews. Prior to the interview, oral or written informed consent was received from all participants, including permission for audio recording. Both individual and focus group interviews followed an open-ended interview guide that was tailored to each community’s unique successes and challenges, based on interview data consolidated over the previous three years (S1. File). Interviews were transcribed by an external transcriptionist and underwent a thorough quality assurance process to ensure accuracy.

Thematic analysis was used to analyze the transcripts and inductively identify themes from the dataset. The primary coder [AC] began with an open-coding process, breaking down ideas and discrete concepts from the text into “codes” (39). Codes were then discussed and agreed upon by the research team and a consensus codebook was developed. This codebook was applied to the dataset by the primary coder using NVivo 12 software. Theme generation was again a shared project within the research team, stemming from discussion of the overarching themes and patterns identified across the final year of interviews. Themes were derived through engagement with the data rather than through the application of an external theoretical framework.

### ThoughtExchange dataset approach

The second dataset contributing to this summative evaluation was collected through a real-time interactive platform, *ThoughtExchange*, during an annual, virtual RSON meeting with clinicians and administrators from the RSON sites. Participants viewed a consent form detailing the use of their response data and were informed that consent was implied if they clicked the link to the ThoughtExchange platform. Participants were given 8 minutes to respond in real time to the perceived gains of RSON at their site (*ThoughtExchange 1,* at the beginning of the meeting) and their view of the interventions most important to continue past the official duration of the initiative (*ThoughtExchange 2*, at the end of the meeting). The platform enabled confidential sharing of information and the prioritization of responses through rating others’ responses out of 5 stars as they were posted. We enabled access to the ThoughtExchange site for an additional 14 days to allow those who were not able to attend the meeting to respond. In total, invitations were extended to approximately 100 potential participants.

Using this interactive feedback tool augmented the more detailed and nuanced understanding gained through interviews and focus groups while respecting the limited time available to clinicians and healthcare administrators in the communities. Taken together, the convergence of these two methods yielded a rich data set revealing the most important outputs of the initiative from the communities’ perspective.

Like with data from the open-ended interviews, data from the two ThoughExchanges was analyzed using a process of thematic analysis (39), and followed a similar approach described above.

### Methodological rigour

Given that researcher interpretation was central to the generation of themes and codes, the practice of reflexive dialogue was incorporated into the analytical process to enhance the rigour of the methodology, with researchers reflecting together on positionality, background, and relationship to the phenomenon to identify potential predispositions or distortions related to the data. Additionally, the analysis was iterative, and researchers returned to the datasets repeatedly throughout the analytical process to ensure the accurate representation of the findings (40). These data are thematically explored, below.

## Results

In response to the first ThoughtExchange, “What were the gains and benefits of RSON at your site?”, 64 respondents provided 93 thoughts and 1249 ratings (S2. File). The second ThoughtExchange was framed by the question, “In an ideal world, which are the most crucial interventions to sustain surgical & obstetrical services at your site?” 60 respondents provided 81 thoughts and 973 ratings (S3. File). In this exchange, respondents frequently linked “crucial interventions” with their perception of the greatest RSON wins seen at their sites over the course of the initiative. The professional designations of respondents to each ThoughtExchanges are reflected in Figures 1 and 2, respectively.

**Fig 1:**
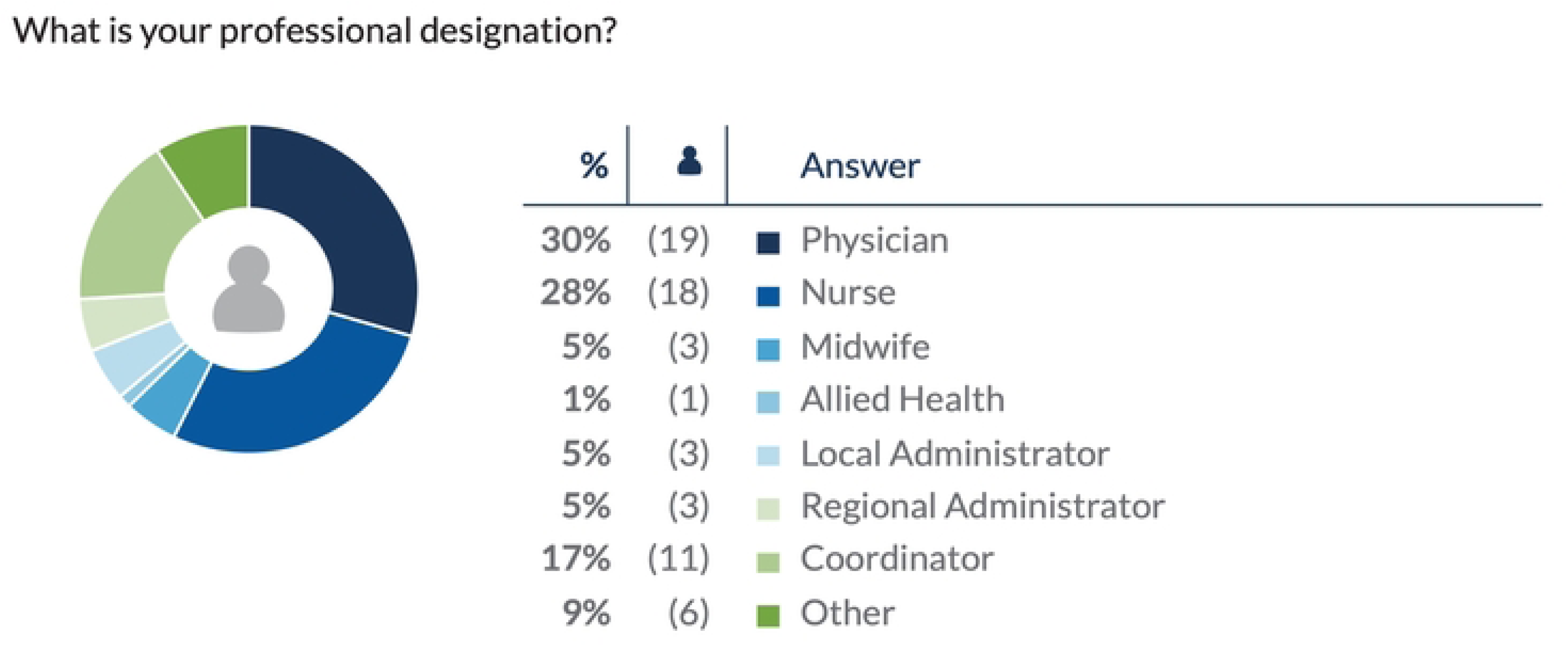
**Professional designation of ThoughtExchange 1 respondents.**

**Fig 2:**
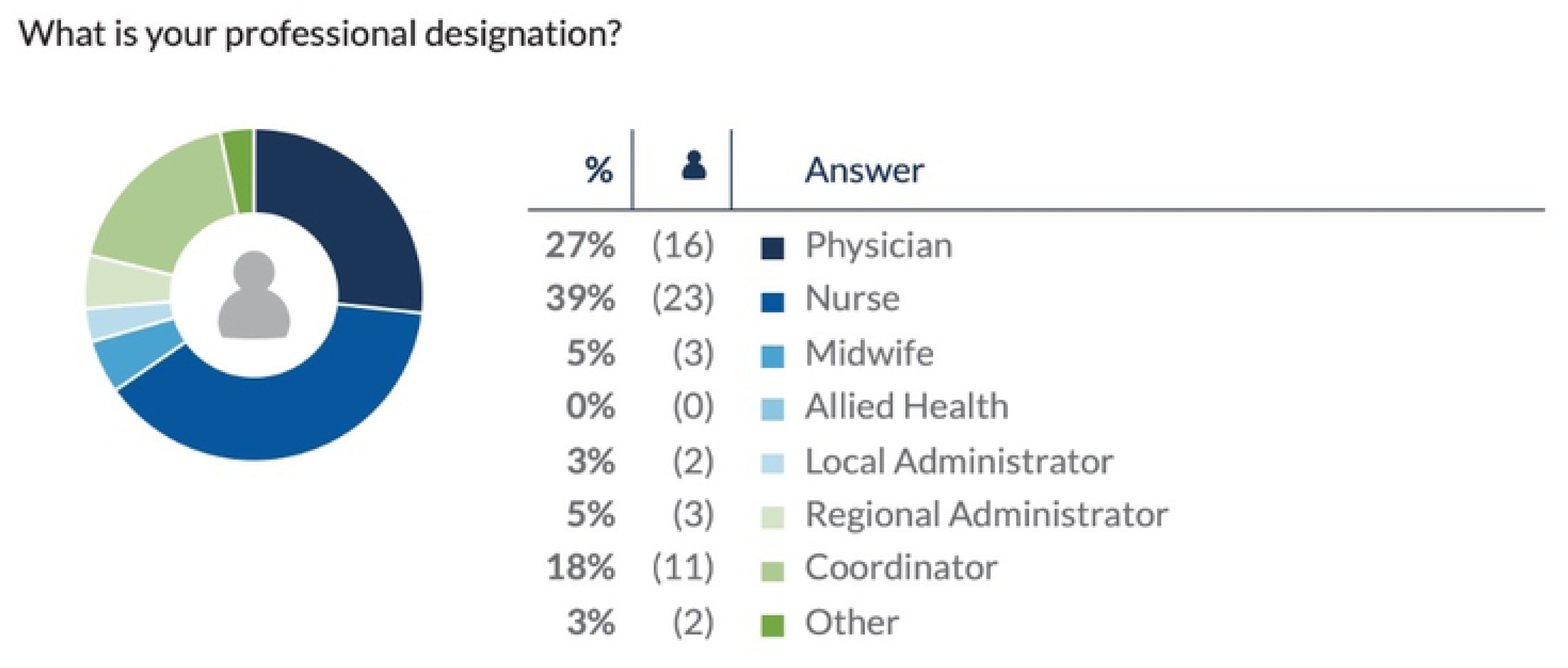
**Professional designation of ThoughtExchange 2 respondents.**

Six overlapping themes emerged from the combined ThoughtExchange dataset. These in turn converged with the findings from the final qualitative individual interviews (n=72) and focus group interviews (n=3) conducted at 7 RSON communities. The professional designations of the qualitative interview participants are represented in Table 1.

**Table 1.** Professional Designation of Interview Participants.

| Professional Designation | Number of Participants |
| --- | --- |
| Physician | 29 |
| Nurse | 30 |
| Midwife | 5 |
| Administrator | 8 |
| Coordinator | 3 |
| Other | 4 |
|  | Total: 79 |

Given the commonalities across all data sources, findings are presented in aggregate.

Congruent thematic areas between the two ThoughtExchanges and the qualitative interviews included *stabilization of local procedural care through increased local scope and volume, ongoing educational supports, quality improvement initiatives, support for team-oriented care, increased administrative structures,* and *achieving appropriate staffing levels.* One additional unique theme, *impact of evaluation data*, was identified during the first ThoughtExchange (‘RSON wins’). Further visualizations of the data as organized by theme are included as supporting information (S4 File).

Themes unique to the summative interviews included those addressing limitations of RSON funding, as the ThoughtExchanges focused mainly on overarching strengths. These themes included *constraints on funding* and *contextual factors limiting participation*. Table 2 summarizes the breakdown of themes by dataset.

**Table 2.**
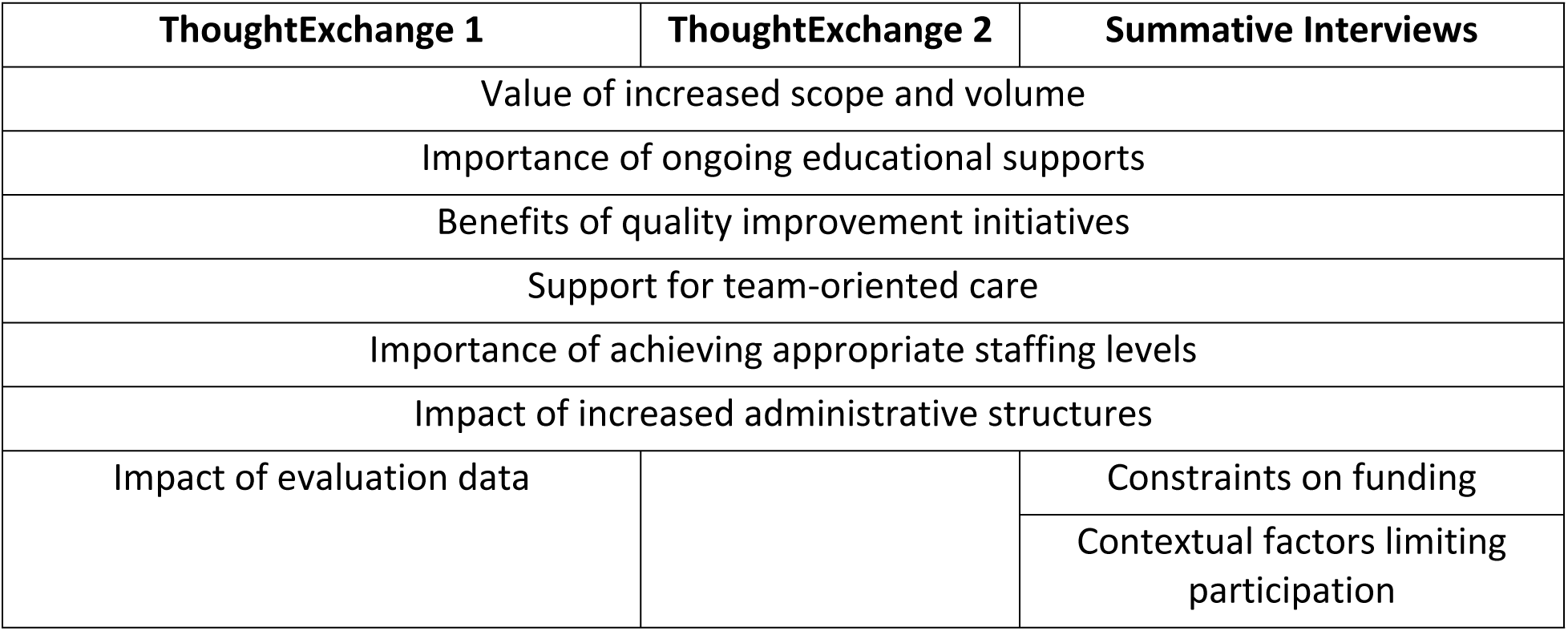
Themes by Data Source.

Across the board, RSON-supported system interventions (pillars) were cited as crucial interventions for service sustainability. One participant summed it up succinctly: “Without RSON, our maternity services would likely have collapsed.” The adaptability and incorporation of local leadership allowed sites to take the resources available through RSON and tailor them to the needs of their communities, which was a departure from an urban-based, one-size-fits-all approach. “It’s made an incredible difference over time,” shared an interview participant, “And it’s not just one thing, it’s all the pillars…that have contributed a little bit. And we built on that over time.”

### Value of increased scope and volume

Increases in scope, the range and type of surgical procedures, and volume of all procedures was identified as both a significant RSON “win” as well as a crucial intervention that supported site sustainability. These augmentations were reported to “create robust program[s]” as well as increase staff retention and engagement. Increased scope of procedures was made possible due to increased outreach services by visiting specialists and augmented local services enabled by coaching and educational support. In practical outputs, sites reported that RSON had precipitated a substantial increase in functional operating room (OR) days, in one instance from 1.5 to 4.5 days per week. Other respondents were impressed by how much busier their operating room had become “simply through some funding and an opportunity to try.”

A key area of increased scope in many sites was orthopedics, with participants reporting opportunities to add several procedures at their site, such as carpel tunnel repair, knee scopes, and ACL repairs. One interview participant shared, “I think that we’re sitting in a really amazing spot right now in terms of the OR specifically. We have a lot of momentum, and we have a lot of days booked coming up for orthopedics. I think it’s amazing that there was growth like this when it was up against so many challenges.” Participants reported that increasing volume also contributed to the sustainability of other services vital to providing surgical care, namely anesthesia. As one respondent explained, “guaranteed work and guaranteed pay” contributes to the sustainability of anesthesia services in rural sites, and one way to achieve these guarantees is to increase scope and volume. Some respondents also noted the value of enhanced volume in tackling waitlists and decreasing the time it takes for patients to receive care.

#### Increased skill development

A byproduct of increased volume at rural sites was the concomitant increase in skill development, as well as positive relationships with colleagues. Respondents found that familiarity with colleagues led to growing competency and confidence in practice, vital in rural, low-volume sites, where repetitions of certain skills may not be as readily available. Increased volume and subsequently increased provider experience was deemed “key to maintenance of healthy and safe OR environments.” As another participant reported, “…repetition has allowed our team to work effectively and efficiently and be better prepared for emergencies.” Additionally, the increased opportunities to “brush up on surgical skills” was of particular importance for new and early-career staff members.

#### Care closer to home

For many, a substantial RSON win was the increased availability of local services, reducing the need for patients to travel out of their community to access care, particularly in the context of obstetrical care. This growth helped prevent service disruption at a time when other services across the province were closing, despite a widespread understanding that, “maternity care close to home is vital to the health of women and communities.”

### Importance of ongoing educational supports

Many participants identified funding for clinical coaching and training as the most crucial intervention to stabilize their sites. Ongoing education was associated with increased provider satisfaction and retention. Several participants observed that participation in education opportunities had been considered a voluntary, “altruistic” task, and the new availability of funding meant that all providers, across professional disciplines, received appropriate compensation for their time. This validation was important to participants, particularly nurses and midwives, for whom funding had not previously been reliably available.

One participant explained that funding also opened doors for clinicians to seek “educational opportunities that might not be feasible to attend” without funding, due to the program costs, but also the costs associated with travel. Respondents suggested that appropriate compensation for education was a protective factor against burnout. Furthermore, funding for multidisciplinary simulations offered teams the chance to review “rare” but critical events from a team-based perspective.

#### Funds for clinical coaching

Participants emphasized the impact of Clinical Coaching in improving service sustainability. Both the availability of local peer coaching and regional specialist coaching was seen as a significant gain at RSON sites, in each instance, supported by remuneration for the coach and coachee. Where there may have previously been hesitancy to reach out to an “off the clock” peer or specialist for support, the formalization of coaching roles and funding streams eased those apprehensions. As one participant explained, “…there’s still a chance you’re interrupting dinner time, but they’re getting paid and so are you. People are less hesitant to phone now. Now nobody’s helping anybody out or doing a favor, right? Now … it’s just work and being valued as such.”

Participants reported that clinical coaching from regional and provincial providers helped them “keep skills fresh”, as well as improve regional care coordiation, and contributed to ensuring positive patient outcomes. It provided opportunities for clinicians to experience higher patient volumes and acuity, either through traveling to regional centres for coaching, or training under a specialist who brought their own caseload to the rural site. Another echoed this sentiment, suggesting that learning through others’ experience is the “only way small communities can maintain high quality standard of care, despite low-volumes, lower opportunities.”

Participants also expressed an increased sense of confidence because of expanded opportunities to practice skills and to develop their professional support networks through mentorship between rural providers and other peers and specialist coaches. One respondent shared, “Mentoring encourages our rural practitioners to build relationships and [gain] comfort in conducting procedure they don’t do often.” Recurrent onsite training and continued opportunities for education played a crucial role in ensuring providers remain confident in their skills and experience cohesion within their teams.

Other participants emphasized the value of interdisciplinary coaching within sites, suggesting that coaching and simulation education created opportunities for “collaborative, interagency work,” leading to improved communication and familiarity within and between sites.

### Dedication to quality improvement initiatives

One of the strongest thematic areas of RSON wins was the formalization of continuous quality improvement (CQI) programs at site levels. Participants reported that the RSON CQI pillar provided structure and created the space for clinicians to specifically protect time for pursuing quality projects and, as a result, improved clinical skills and enhanced patient care. For many, this came down to establishing a “culture of learning and embedded quality improvement” at their sites.

Projects such as needs assessments for nursing teams and tracking tools which help chart the “decision to incision” process for cesarean sections were co-created with local sites, and provided clinicians and administrators the data to see in what areas improvements would be most helpful. Having survey feedback meant that teams could “objectively assess outcomes” and tailor processes at their sites to best address specific limitations. Respondents noted that CQI initiatives were also used to modernize policies, helping to ensure that teams were up to date on best practices and making the most efficient use of time and resources.

#### Local autonomy

Participants shared that a specific benefit of the RSON CQI pillar was its local implementation and management. Some participants felt that generic quality improvement initiatives were often developed with larger urban sites in mind. In contrast, having direct access to funding and tools for CQI meant that RSON teams had the autonomy to develop their own initiatives in response to local (rural) needs. This independence and control over QI funding meant that facilities could “level up their care by focusing on what’s important to them.” Participants also reported quicker project turnarounds and an appreciation for the “agility” they had to adjust initiatives in real time if they were not meeting site-level needs.

Autonomy over funds meant less “administrative hassle,” leading to more uptake and local staff buy-in. Others discussed how crucial it was to have designated budgets for quality improvement initiatives, such that financial resources are not “swallowed up” by other budgets. One participant explained, “…having a nimble little local committee to create change” was much more effective and “user-friendly” than instituting a top-down, one-size fits all model dictated by external bureaucracy.

> *I do think that that’s what the appeal of RSON was … that it acknowledged that it’s not a formulaic approach for every hospital and site. And that it was self-directed. I think a lot of the momentum came from the acknowledgement and autonomy to make those decisions.*

#### CQI nurse position

Along with the time and structure the CQI pillar provided, participants also affirmed the value of the CQI nurse position funded through RSON. Respondents appreciated that the position meant someone was working to “address deficiencies that no one else previously had responsibility for.” The position was vital to ensuring that education and quality initiatives were not slipping through the cracks due to pressing clinical demands.

Finally, participants found that funding for RSON CQI and the CQI nurse role augmented their “networking capacity” through establishing avenues for collaboration with other networked sites. Participants appreciated the opportunities they had to communicate about quality initiatives with colleagues across the province at RSON communities, to see what was working well at their sites and get new ideas for projects.

### Support for team-oriented care

Participants repeatedly identified team-oriented work as a benefit of RSON at their sites and a crucial intervention for sustaining their services. Improved sustainability concomitantly improved recruitment and retention efforts as participants reported that “sustainability is easier to recruit into…”

RSON was found to promote strong leadership, help create a “cohesive team dynamic,” and strengthen collaboration between providers. Participants valued opportunities to learn and practice as a team, improve their communication and team culture, and develop mechanisms of positive support and skill building.

#### Interdisciplinary collaboration

Participants discussed several approaches to supporting team-oriented care, including cultivating opportunities for interdisciplinary relationships, integrating siloed teams (such as surgery and maternity), and improving team communication and collaboration. Participants found that funding for RSON working group meetings created an environment where nurses, midwives, and physicians were consciously working side-by-side and towards the common goal of improving program sustainability. RSON resources meant that everyone was remunerated for their time when participating at these meetings. RSON-related working groups and training opportunities were credited with an increase in communication between team members and an organic development of cross-disciplinary respect. As one participant explained, “[RSON] created a sense of community and camaraderie and made us so much more resilient when COVID hit our teams. We’re so strong because we supported each other.” Another respondent noted, “…it’s no longer about surgeons booking slates and nurses turning up to shifts. There’s a lot more teamwork going on than that.”

One respondent explained the benefits of working as a high functioning team: “You … have better communication, someone to provide guidance or to develop a plan with, and overall, more efficient, professional care [for] our patients.” For this participant, and others, support for team-based care was one of the most significant interventions to sustain surgical and obstetrical services at their site.

#### Regional relationships

The development of relationships between and across teams extended beyond local teams to the regional level. RSON provided the administrative capacity to support regional and provincial networking between rural sites and regional referral centres, enhancing referral specialists’ knowledge of the work being done at rural sites and bolstering their confidence in the care provided by rural clinicians.

> *To think that I could have not left my tiny little community…for two years, but I still have met tons of people all across the province in all different kinds of provider roles, it’s really neat and I think that RSON has done a fantastic job of the networking aspect.*

#### Health authority relationships

For some, RSON offered groundwork for building more substantial relationships between rural sites and regional Health Authorities. These relationships were seen as essential for ongoing site sustainability.

Participants also recognized the value of RSON in providing small sites a seat at the table and allowing them to influence higher level decision making. Some participants voiced concern that the end of RSON would mean the loss of this communication pathway. One shared, “That would be my biggest fear…that with the end of RSON funding, that we lack the ability to affect change at a higher level, or to be listened to.” Accordingly, participants voiced a need for ongoing supports to maintain their capacity to institute change at regional levels, communicate with decision-makers, and advocate for their needs.

### Achieving appropriate staffing levels

A thematic response about the interventions necessary to sustain services was ensuring appropriate staffing levels at rural sites. Respondents were clear that without adequate staffing levels, providers felt unsupported, which not only impacted morale and retention, but also the provision of safe patient care. This is further explicated below.

#### Funding for additional staffing

Additional staffing afforded by RSON was a benefit noted by many respondents. Others shared how having consistent and stable baseline staffing “creates capacity for things beyond crisis management.” Rather than expending all resources trying to stay afloat, sites with appropriate staffing levels were described as having the time, resources, and energy to engage in further sustainability interventions, such as ongoing professional education and quality work.

Participants also discussed the impact of understaffing on the provision of services themselves. Issues such as inconsistent surgical back-up for the OR, due to gaps in call coverage, were connected to OR closures, a threat to overall sustainability.

#### Improved provider well-being

Having more staff available meant increased support for new staff who sought mentorship and guidance, and coverage for nurses who previously were unable to take their vacation time without leaving the OR short staffed. Both factors contributed to reducing fatigue and burnout. As one interviewee noted, “We used to just work very minimally [staffed] and not get any breaks and then we got the new funding, and it was just a dream.” Ultimately, participants reported that funding for staffing increased provider retention, job satisfaction, staff morale and sustainability as the work was spread more consistently across more individuals. Another interview participant shared, “There’s a lot more people willing to take on the work because they’re not overwhelmed.”

#### Nursing staffing

Many participants noted the specific importance of optimizing maternity and OR nursing staff levels, reiterating that nursing is fundamental to site stabilization. As one respondent wrote, “We cannot deliver our patients here if [maternity nurses] are not here, feeling safe and supported at work.” Another shared that appropriate nursing levels increase “safety, retention, [and] provider confidence.” Respondents also discussed the beneficial effects of being able to hire “regular” nursing lines rather than relying on casual positions which did not contribute as much of a sense of stability.

### Impact of increased administrative structures

Respondents identified the *structure* of the RSON program as a key stabilizing element of their services. This included administrative infrastructure and positions, such as local and regional coordinator roles, CQI leads (often a Registered Nurse), and formalized working groups, which provided forums for staff, physicians, and nurses to connect and discuss how to achieve surgical sustainability. These administrative structures allowed sites to build their “capacity to carry out the work” through developing local leaders responsible for organizing projects and further interventions. One participant discussed the significant impact of their local working group, saying they “keep us connected and working towards the same goals.” Having access to a larger network of support, both clinical and research related, was also identified as a significant benefit.

Respondents particularly emphasized the central role of local community coordinators (LCCs) in managing RSON funding. Participants discussed the importance of coordinators in knowing the nuances of the site and being able to work with teams and existing initiatives in order to effectively assign funding. As one participant explained, “coordinators…put all the pieces of generalism together on the ground to help the teams to work clinically.” Participants also highlighted the cost benefits of LCCs, finding their work “invaluable” and a relatively “cheap resource” compared to having administrative and organizational roles fall to clinicians, or become “scattered” or lost all together.

Participants also described the significant impact of having established, centralized RSON infrastructure at a regional level, which enabled “cross provincial ties” between RSON sites. Participants found this led to a “rural cohesive approach to patient care,” something they identified as vital to sustaining surgical and maternity services at their sites and across the province. Pan-provincial RSON administrative and leadership team were identified as vital to supporting “data collection, equitable access to care, connection to rural provincial partners, and standardization [of] maternity services.”

### Impact and use of evaluation data

One final outstanding RSON win was related to the value of the RSON evaluation itself, with participants voicing a strong appreciation for evidence-based feedback on the efficacy of their sites. Participants were invested in receiving “accurate and easily accessible data” to help them optimize their programs, demonstrate the credibility of their small surgical sites, and showcase safety and quality of their work to larger referral sites and governing bodies. Respondents shared that site-level findings provided them some concrete validity in asking for further support from the Ministry of Health and health authorities, given the demonstrated return on investment. As one noted,

> *We have worked hard on documenting all the projects and initiatives and quality improvements that we have been working on, with the idea that when RSON ends, we can show hard data to the [Health] Authority about…listen, if we’re not provided with the same kind of support, all of this is going to not happen.*

Alongside RSON wins, participants also shared their experiences of limitations within the RSON program, which centred around two general themes: experiences of funding constraints, and factors which reduced participation in RSON.

### Funding constraints

#### Lack of local autonomy in funding usage

Though most participants reported positive experiences with the level of local flexibility granted in using RSON funds, some felt constrained by a lack of sufficient autonomy in how funding was allocated. This feedback centred around perceived “tiers of funding,” where participants understood there to be funds allocated to their local site, as well as a larger pool of RSON funding under the direction of health authority decision-makers. Some stakeholders voiced frustration that the “big money,” (higher level scope and volume funding), was controlled by the health authority and spread between communities at their discretion. One participant shared that their working group “didn’t have a lot of say and influence in what was actually happening there.” This participant suggested that though RSON leaders from their local community were integrated into decision-making tables at a high level, as the project evolved, “it’s still not as site specific or community driven as it first appeared.” Others expressed some confusion over how these tiers of funding integrated and a desire for more grassroots-level involvement.

#### The regional/local divide

Some participants expressed concerns regarding which level (regional or local) of the healthcare system was benefitting from RSON funding, identifying a historic sense at some rural sites of being neglected at the provincial level and consequently, concerns of being overlooked by decision makers allocating RSON funding. Participants desired increased communication between sites and clarity on how funds distributed to larger referral centres were also benefiting their local site.

#### Lack of clarity around budgets

Another limitation expressed was a sense of confusion around what exactly the RSON budget had been used for and how to access funds. Most participants who provided this feedback acknowledged this was due to their own limited involvement with the program, but nevertheless, a desire for increased transparency with site members at all levels of involvement with RSON was noted.

#### Ineligible funding opportunities

Participants did report some limitations as to what RSON funds could be used for, with one local site coordinator sharing the challenge of being asked for “larger capital things” such as funding for increased local scope and volume, and having to tell colleagues that doesn’t fall under their budget. Others discussed the limits of funding for specific pillars which couldn’t be reallocated for other purposes deemed higher priorities at each site. For example, while participants understood why many equipment upgrades fell outside of the scope of RSON funding, several still identified a significant need for new equipment in their communities, and a growing imperative to find funding from other sources to meet these needs.

#### Promising program, precarious funding

A few participants touched on how the original time-limited nature of RSON funding, (5 years), limited their investment in the initiative. Some participants stressed the importance of knowing ahead of time whether any funding streams would be taken up by the health authorities or other grants, and if any RSON funded positions could remain. Concerns around succession planning were prevalent in participants’ feedback. One explained,

> *I suppose we’re at the point where we’re starting to worry about when RSON stops. And that maybe says how well it’s been helping us. It does help us fund a lot of things that allow the physicians to get together and to get together with a team of nurses and other providers. And when that falls away, we might have difficulties, because then you’re not problem solving on a regular basis.*

Succession planning was of particular importance to sites who joined the RSON program after the start of the five years: “I really hope that more funding is attainable, and that it can continue, because we’re just getting started. So, I would love to see it continue and actually make those changes.”

### Contextual factors limiting participation

Another reported limitation was the buy-in and time commitment required from sites to make best use of RSON resources and to see certain advancements made. Most cases where participants declined involvement with the RSON project could be traced to an overall lack of personal and professional capacity to participate in *any* initiative outside of clinical workloads.

RSON projects were seen in some cases to add to providers’ workloads in impractical ways, especially at sites where staffing was most precarious. One participant explained, “It was tricky to adjust to the mindset of what we would think of as extra work, like the CQI portion of it all.” While many participants maintained that CQI projects were among the most valuable aspects of the RSON project overall, others found that the time and energy investment associated with these projects was a cost to participation that they could not afford.

Another participant described the challenge of having multiple support initiatives at sites, fearing that providers would burn out on opportunities and end up withdrawing from helpful programs. This idea was compounded by the suggestion that sites were often “calling upon the same players” to support and lead various projects, including RSON.

## Discussion

Locally available surgical and procedural care in rural communities not only increases access to care for the local population, but also stabilizes other health services such as urgent and emergent care, local intrapartum care, and screening and diagnostics (41). To achieve this, however, health planners must contend with staffing shortages (42–45), lack of investment in infrastructure (43,46) and, at times, challenging relationships between rural proceduralist and regional specialists (47). New models of health system organization and funding are needed to respond to these challenges; such innovation was enabled through the structure and values propositions of RSON funding.

RSON funding addressed some of the gaps that make participation in skill development, education, and local quality improvement initiatives difficult due to un-funded time away from clinical and administrative responsibilities. This up-stream approach addressed, in part, the tension between the recognized need for these activities and the loss of income incurred in a fee-for-service billing system by tacitly acknowledging the need for discrete, focused funding streams for individual practitioners and community teams. Almost all participants expressed appreciation beyond the actual funding for the proxy for respect that this funding provided.

The innovation inherent in the RSON funding model gave rise to secondary benefits, beyond infrastructure support, such as the enhanced regional relationship catalyzed by clinical coaching, relationships which are essential in a well-functioning healthcare system. Outside of the immediate clinical encounter, buttressed regional relationships led to the reciprocal sharing of resources, including RSON providers offering care in regional hospitals to offset staffing shortages (4). Ultimately, enhanced relationships allow for a unified rural voice in advocacy for policy change to support challenges around workforce development and ultimately health equity.

Ultimately, the stabilization of low-volume, generalist procedural care is contingent on having enough procedural scope and volume to ensure efficiency and effectiveness (patient safety), which is sometimes challenging to achieve due to limited funding resources. Beyond meeting a broader range of the immediate healthcare needs of local patients, funding for increased scope and volume precipitated responsiveness to locally prioritized procedures, for example, orthopedic surgery in communities proximal to ski resorts. As this was undertaken through a regional – and provincial – lens, resources *between* communities could be optimized, avoiding oversaturation of specialized procedures across low population densities. In turn, the increased volume of procedures that additional health human and capital resource funding allowed led to higher resource utilization due to economies of scale (fixed costs spread over a larger number of procedures). Likewise, increased procedural volume resulted for many RSON participants in increased confidence with certain procedures, which both enhances quality of care and provider sustainability.

The limitations in the RSON initiative noted by participants in the summative evaluation reflected overall health system challenges, namely funding constraints for capital costs and more significant human resource investments. These issues are entrenched within healthcare systems globally and require a system level solution, through structural healthcare reforms. Within healthcare delivery, prioritization of healthcare spending to reflect the greatest needs (such as primary and preventative care) for the most underserved population is essential.

Along with participants’ valuation of discrete activities enabled by RSON that provided benefit to health service delivery and sustainability in their communities, was the tacit allusion to the innovation of the network structure itself. That is, many of the activities that were noted positively in the evaluation – such as increased communication with regional referral specialists, the capacity to learn from successes in other rural communities, and increased teamwork at a local level – were outputs of the network structure. Implementing regional networks of surgical and maternity care was the cornerstone of RSON’s vision; doing this with regard for local autonomy and facilitating learning through centralized planning enabled the RSON initiative to actualize this vision.

Findings from this evaluation point to universal challenges of sustaining rural health services in rural settings and reveal strategies that may be adapted to meet local needs in other jurisdictions, particularly those where small hospitals are under the threat of closure or service reduction. Universal themes include the importance of local autonomy in resource allocation and priority-setting, in resistance to urban designed policies not applicable to rural sites (48). As we are currently experiencing a global health human workforce shortage (49), the demonstration of the efficacy of up-stream investment (clinical coaching, local and regional team development) is supported, reflecting the growing evidence of attrition when rural providers are under-supported and isolated (50). Likewise, participant response to the value of localized means to determine priorities, collect, analyze and apply local data speaks to the value of local data in a ‘data aggregated’ health system, where rural concerns are often overshadowed.

Perhaps most importantly, findings from the wider evaluation, reinforced by participant narratives through the summative evaluation, attest to a counter-narrative to the contention that low procedural volume leads to poorer health outcomes. Participants – and outcomes data from this study (51) – demonstrated that when well supported, low volume sites can offer high quality care while meeting the mandate of “closer to home”, which is a structural reality of care across many jurisdictions internationally. The nuances of how resources are offered to support these sites, such as limitations of time-limited funding and the importance of supporting local autonomy to resource allocation decisions, apply equally to other settings.

## Limitations

Although data for this summative evaluation was triangulated from qualitative interviews and real-time participant input through a virtual platform exchange, which increased the rigour and reliability of findings, the following limitations are noted. Due to the summative nature of the evaluation and collection of end-of-study data, participant recall bias may have occurred, highlighting the most recent or impactful events. In addition, the consistent participant cohort through the data collection cycle may have introduce representation bias, where those with more critical opinions tended not to engage. Limitations to the use of the ThoughtExchange platform included the brief window for input of comments and responses (8 minutes), which may have precluded more in depth and thoughtful considerations. However, we feel this was mitigated by the inclusion of rich interview data.

Data from this study is context-specific to the political and pragmatic organization of health services delivery in British Columbia, and all findings may not be applicable to other jurisdictions with variant contexts. Finally, although RSON was a longitudinal initiative, interviews for the summative evaluation were captured in the final year and are not able to demonstrate changes over time.

## Conclusion

Findings from the summative evaluation of the RSON initiative alerted us to the importance of a *wrap-around* approach to health service interventions. This need reflects the complexity of health service delivery and the imperative to coordinate different aspects of stabilizing interventions to ensure they have a supportive context and work in synergy with other initiatives. Beyond stabilizing services, a comprehensive approach is better poised to support innovation, as it mitigates system stressors from different perspectives. In the RSON initiative, this allowed site teams to be responsive and agile in addressing local needs as they emerged, in a way that meaningfully involved the interdisciplinary hospital team. A coordinated, holistic approach to stabilizing rural surgical and obstetrical services in the study jurisdiction was the overarching intent – and success – of the RSON initiative. Additionally, more discrete benefits were noted from community members receiving RSON interventions, namely the *infrastructural advantage* of having community-based coordination for the funding (and often other funding as well to reduce unproductive redundancies) and a point-person for quality initiatives. Additional value was attributed simply to the focus that the funding put on local services or, as one participant said, “valuing the work [we] do, which isn’t often seen.”

Despite structural health service advantages, perhaps the most important output of RSON was the *cultural change* in key determinants of stability and at individual site levels, a necessary precursor to long-lasted change. The combination of this retrospective assessment of RSON by those directly involved and the program evaluation findings noted above provide a strongly defendable position to argue for continuation and expansion of program funding.

## Data Availability

All relevant data from the ThoughtExchange dataset is contained within the manuscript and Supporting Information Files S2 and S3. Qualitative data from the larger RSON interview and focus group dataset cannot be made available on a public repository because participants did not consent to this use of their data. The text data contain sensitive, potentially attributable information. These restrictions are in place in accordance with the UBC Behavioural Research Ethics Board minimal risk agreement. Future interested researchers may contact their institutional ethics review board or UBC Ethics Board to request access to confidential data: https://ethics.research.ubc.ca/about-human-research-ethics/contact-us#ubc-breb.

## Acknowledgements

We extend our gratitude to Kathrin Stoll, evaluation coordinator, and to Simrat Dial for support with manuscript preparation. We thank Payal Parti for her contributions to the background literature review. We are forever grateful to the participants at each site who shared their experiences with us. The authors would like to express their deep appreciation to our funders, BC’s Joint Standing Committee on Rural Issues.

## Supporting information

**S1 File. Interview guide**

**S2 File. ThoughtExchange question 1 responses.**

**S3 File. ThoughtExchange question 2 responses.**

**S4 File. Data Visualizations.**

**S5 File. COREQ (COnsolidated criteria for REporting Qualitative research) Checklist**

## Notes

### Competing Interest Statement

Tom Skinner was employed by the Rural Coordination Centre of BC (RCCbc) as the Project Manager of the RSON Initiative. As an employee, his travel to the rural communities was covered because he was supporting hospital teams in implementing the PROES survey and interpreting the result. All other authors are part of the RSON evaluation team at the University of British Columbia, Canada and have no conflicts of interest to declare. This does not alter our adherence to PLOS ONE policies on sharing data and materials.

### Funding Statement

Yes

### Author Declarations

This study received approval from the Behavioural Research Ethics Board of the University of British Columbia (BREB H18-01940). All qualitative interview participants provided recorded, verbal informed consent to participate before each interview, and each was educated on their right to withdraw their participation at any time. The University of British Columbia Behavioural Research Ethics Board approves of recorded verbal informed consent. Participants from the ThoughtExchange dataset viewed a consent form prior to accessing the platform, which detailed use of their response data. They were told that consent to participate in the study was implied by clicking the link that directed them to the ThoughtExchange platform at the bottom of the consent form page. This consent form and process of obtaining informed consent was approved by the UBC Behavioural research Ethics Board (BREB H18-01940).

